# The burden of isolation to the individual: a comparison between isolation for COVID-19 and for other influenza-like illnesses in Japan

**DOI:** 10.1101/2021.08.19.21262267

**Authors:** Shinya Tsuzuki, Norio Ohmagari, Philippe Beutels

## Abstract

At present, there is scarce evidence about how much burden the isolation of COVID-19 patients is. We aimed to assess the differences between COVID-19 and other influenza like illnesses in disease burden brought by isolation. We conducted an online questionnaire survey of 302 people who had COVID-19 or other influenza-like illnesses (ILIs) and compared the burden of isolation due to sickness with one-to-one propensity score matching. The primary outcomes are the duration and productivity losses of isolation, the secondary outcome is quality of life (QOL) valuation on the day of the survey. Acute symptoms of outpatient COVID-19 and other ILIs lasted 17 (interquartile range [IQR] 9-32) and 7 (IQR 4-10) days, respectively. The length of isolation due to COVID-19 was 18 (IQR 10-33) days and that due to other ILIs was 7 (IQR 4-11) days, respectively. The monetary productivity loss of isolation due to COVID-19 was 1424.3 (IQR 825.6-2545.5) USD and that due to other ILIs was 606.1 (IQR 297.0-1090.9) USD, respectively. QOL at the time of the survey was lower in the COVID-19 group than in the “other ILIs” group (0.89 and 0.96, *p* = 0.001). COVID-19 infection imposes a substantial disease burden, even in patients with non-severe disease. This burden is larger for COVID-19 than other ILIs, mainly because the required isolation period is longer.

## Introduction

Coronavirus disease 2019 (COVID-19) caused by the SARS-CoV-2 virus, has become a global health threat [1–3]. More than half a year after the roll-out of highly effective vaccines against COVOD-19, the pandemic remains difficult to control [4].

COVID-19 can be regarded as one of the influenza-like illnesses (ILIs) because it causes upper respiratory symptoms like seasonal influenza and its severity is mild in most cases [5–7]. However, there are several distinctive characteristics which differentiate COVID-19 from other ILIs.

First, COVID-19 is more likely to result in severe illness and death than other ILIs do. Previous studies suggested that the infection-fatality rate of COVID-19 was about ten times higher than that of seasonal influenza [5,8]. Although patients often recover with only mild symptoms, COVID-19 infection should be another important cause of excess death [9].

Second, the transmission dynamics of COVID-19 is quite different from other ILIs, in that pre- and asymptomatic transmission is more common than for other ILIs [10–12]. Additionally, both incubation period and infectious period of COVID-19 were regarded as longer than those of other ILIs [13,14]. These facts suggest that COVID-19 required a longer duration of isolation as a countermeasure in order to slow down its spread.

Presumably, we need to strengthen our isolation policy in order to prevent further spreading of COVID-19 due to the reasons mentioned above. However, at the same time, we have to consider the societal burden of such intervention including productivity loss because its appropriate duration should be determined by a kind of trade-off between disease prevention effect and societal loss.

At the early stage of the pandemic, the Japanese government determined the duration of isolation for COVID-19 patients as two weeks [15], following the recommendation published by World Health Organization (WHO), Centers for Disease Control and Prevention (CDC) and other organizations [16–18]. This recommendation was updated as “10 days after symptom onset, plus at least 3 additional days without symptoms” in June 2020 [15], based on the updated recommendation published by WHO and other organizations [16–18].

However, these recommendations were defined based on clinical insights. It would be relevant to try and define the duration of isolation with both clinical, economic and societal aspects taken into consideration because such long duration of isolation may present a substantial burden for the patients.

At present, there is scarce evidence about how much burden the isolation of COVID-19 patients is. The main objective of the present study is to estimate its burden from a societal perspective.

## Methods

### Settings

We conducted an online questionnaire survey of 302 people who had COVID-19 or other influenza-like illnesses (ILIs) between 1^st^ October 2019 and 28^th^ February 2021. The participants were voluntarily and randomly recruited from registrants of NEO MARKETING INC, a Japanese marketing research company. The participants were asked to provide information on the latest episode of isolation due to having ILIs during the study period and stratified them into (a) COVID-19 group and (b) other ILIs group. We defined ILIs as these diseases diagnosed by physicians; COVID-19, seasonal influenza, adenovirus infection, respiratory syncytial virus (RSV) infection, hand, foot and mouth disease, pertussis, and other common colds. We did not use the definition of ILI [19] because asymptomatic infection is common in COVID-19 and other ILIs, in which case not symptoms, but diagnoses provide the rationale for isolation. The responders had to be at least 20 years old. Informed consent was given before the start of the survey.

### Statistical analysis

We collected data on sex, age, number of household members, education level, household and individual income, duration of symptoms and isolation, and Quality of Life (QOL) at the time of questionnaire survey. As for QOL, we used the 15-D questionnaire [20] to estimate QOL value at the time the survey was conducted. Categorical variables were presented as absolute number and percentage, continuous variables as median and interquartile range (IQR).

We compared the COVID-19 group with the “other ILIs” group. The primary outcome is duration and productivity loss of isolation evaluated as monetary value; the secondary outcome is QOL value. We compared the outcomes between two groups using Mann-Whitney U test with one-to-one, nearest neighbour propensity score matching (caliper = 0.2) [21]. Age, sex, education level and presence of underlying medical conditions were adjusted. Monetary value of productivity loss was calculated by multiplication of duration (days) and wage of each participant (per day equivalent). We transformed Japanese Yen (JPY) to USD as 110 JPY = 1 USD, according to the exchange rate in 2021. The QOL values were compared after excluding participants who presented any symptoms at the time of questionnaire survey. Additionally, we compared QOL values between participants who presented any symptoms due to COVID-19 and those do not.

Two-sided *p* values of < 0.05 were considered to show statistical significance. All analyses were conducted by R, version 4.0.5.[22]

### Ethics approval

This study was approved by the Ethics Committee of National Center for Global Health and Medicine (NCGM-G-004001-01).

## Results

Table 1 shows the demographic characteristics of participants. 138 were classified into COVID-19 group and 164 were other ILIs group. The basic characteristics of two groups were similar. The median duration of symptoms and isolation for other ILIs group were 7 days, while those for COVID-19 group were 15.5 and 17.0 days.

**Table 1.** Demographic characteristics of the participants.

|  | COVID-19 group | Other ILIs group |
| --- | --- | --- |
| **Number of participants** | 138 | 164 |
| **Male** | 97 (70.3) | 110 (67.1) |
| **Age (median [IQR])** | 45.5 [37.0, 55.0] | 46.0 [36.0, 56.0] |
| **Number of household members** |  |  |
| **1** | 33 (23.9) | 35 (21.3) |
| **2** | 23 (16.7) | 29 (17.7) |
| **3** | 35 (25.4) | 49 (29.9) |
| **> 4** | 47 (34.1) | 51 (31.1) |
| **Diagnosis** |  |  |
| **COVID-19** | 138 (100.0) | 0 (0.0) |
| **Seasonal influenza** | 0 (0.0) | 122 (74.4) |
| **RSV infection** | 0 (0.0) | 1 (0.6) |
| **Hand, foot, mouth disease** | 0 (0.0) | 1 (0.6) |
| **Pertussis** | 0 (0.0) | 1 (0.6) |
| **Common cold** | 0 (0.0) | 39 (23.8) |
| **Comorbidities** |  |  |
| **Asthma** | 37 (26.8) | 30 (18.3) |
| **Allergic rhinorrhea** | 46 (33.3) | 45 (27.4) |
| **Atopic dermatitis** | 25 (18.1) | 20 (12.2) |
| **Neurological disorder** | 11 (8.0) | 3 (1.8) |
| **Respiratory diseases** | 16 (11.6) | 5 (3.0) |
| **Cardiovascular diseases** | 17 (12.3) | 7 (4.3) |
| **Diabetes mellitus** | 23 (16.7) | 8 (4.9) |
| **Renal diseases** | 16 (11.6) | 7 (4.3) |
| **Liver diseases** | 10 (7.2) | 8 (4.9) |
| **Metabolic diseases** | 13 (9.4) | 2 (1.2) |
| **Immunodeficiency** | 14 (10.1) | 4 (2.4) |
| **Pregnancy** | 4 (2.9) | 0 (0.0) |
| **No comorbidities** | 58 (42.0) | 72 (43.9) |
| **Place of isolation** |  |  |
| **Home** | 86 (62.3) | 157 (95.7) |
| **Hotel** | 51 (37.0) | 12 (7.3) |
| **Hospital** | 51 (37.0) | 7 (4.3) |
| **Duration of work restriction** | 14 [10, 23] | 5 [4, 10] |
| **Monthly wage (USD)** | 2272.7 [1818.2, 4545.5] | 2272.7 [1363.6, 3636.4] |
| **Proportion of wage covered during isolation** | 80.0 [50.0, 100.0] | 95.0 [50.0, 100.0] |
| **Education level** |  |  |
| **Secondary school** | 0 (0.0) | 3 (1.8) |
| **High school** | 22 (15.9) | 42 (25.6) |
| **Vocational school** | 29 (21.0) | 15 (9.2) |
| **University** | 71 (51.4) | 91 (55.5) |
| **Graduate school** | 16 (11.6) | 13 (7.9) |
| **Quality of Life** | 0.89 [0.73, 0.97] | 0.95 [0.84, 0.99] |
| **Duration of isolation** | 15.5 [11, 25] | 7 [5, 12] |
| **Duration of symptoms** | 17 [9, 32] | 7 [4, 10] |
| **Ongoing symptoms** | 11 (8.0) | 1 (0.6) |

ILI; influenza-like illness, RSV; respiratory syncytial virus, USD; United States Dollar
(110 Japanese Yen = 1 USD)
Numbers with brackets represent absolute number and percentage
Numbers with square brackets represent median value and interquartile range

Table 2 describes the details of data after propensity score matching. Standardized mean difference (SMD) larger than 0.1 was regarded as significant imbalance.

**Table 2.** Demographic characteristics of data after propensity score matching.

|  | COVID-19 group | Other ILIs group | SMD |
| --- | --- | --- | --- |
| **Number of participants** | 128 | 128 |  |
| **Male** | 92 (71.9) | 89 (69.5) | 0.052 |
| **Age (median [IQR])** | 44.0 [37.0, 54.0] | 46.0 [36.0, 55.0] | 0.019 |
| **Diagnosis** |  |  |  |
| **COVID-19** | 128 (100.0) | 0 (0.0) |  |
| **Seasonal influenza** | 0 (0.0) | 94 (73.4) |  |
| **RSV infection** | 0 (0.0) | 1 (0.8) |  |
| **Hand, foot, mouth disease** | 0 (0.0) | 1 (0.8) |  |
| **Pertussis** | 0 (0.0) | 1 (0.8) |  |
| **Common cold** | 0 (0.0) | 31 (24.2) |  |
| **Comorbidities** |  |  |  |
| **Asthma** | 36 (28.1) | 24 (18.3) |  |
| **Allergic rhinorrhea** | 44 (34.4) | 38 (29.7) |  |
| **Atopic dermatitis** | 25 (19.5) | 15 (11.7) |  |
| **Neurological disorder** | 2 (1.6) | 11 (8.6) |  |
| **Respiratory diseases** | 15 (11.7) | 3 (2.3) |  |
| **Cardiovascular diseases** | 13 (10.2) | 4 (3.1) |  |
| **Diabetes mellitus** | 22 (17.2) | 5 (3.9) |  |
| **Renal diseases** | 15 (11.7) | 4 (3.1) |  |
| **Liver diseases** | 9 (7.0) | 6 (4.7) |  |
| **Metabolic diseases** | 12 (9.4) | 2 (1.6) |  |
| **Immunodeficiency** | 13 (10.2) | 2 (1.6) |  |
| **Pregnancy** | 3 (2.3) | 0 (0.0) |  |
| **No comorbidities** | 53 (41.4) | 55 (43.0) | 0.0156 |
| **Place of isolation** |  |  |  |
| **Home** | 79 (61.7) | 123 (96.1) |  |
| **Hotel** | 48 (37.5) | 11 (8.6) |  |
| **Hospital** | 47 (36.7) | 74(3.1) |  |
| **Education level** |  |  | 0.019 |
| **Secondary school** | 0 (0.0) | 0 (0.0) |  |
| **High school** | 27 (21.1) | 21 (16.4) |  |
| **Vocational school** | 27 (21.1) | 12 (9.4) |  |
| **University** | 66 (51.6) | 80 (62.5) |  |
| **Graduate school** | 14 (10.9) | 9 (7.0) |  |

ILI; influenza-like illness, SMD; standardized mean difference RSV; respiratory
syncytial virus, USD; United States Dollar (110 Japanese Yen = 1 USD)
Numbers with brackets represent absolute number and percentage
Numbers with square brackets represent median value and interquartile range

Table 3 shows the outcome comparison between the COVID-19 group and the “other ILIs” group. The COVID-19 group showed longer duration of symptoms (18 days versus 7 days) and isolation (16 days versus 7 days). Productivity loss of isolation due to COVID-19 was greater than that due to other ILIs (1424.3 USD and 606.1 USD, respectively). The QOL value of COVID-19 group was lower than that of other ILIs group (0.96 and 0.89, respectively).

**Table 3.** Comparison of outcomes between two groups by matched data.

|  | COVID-19 group | Other ILIs group | *p* value* |
| --- | --- | --- | --- |
| **Duration of symptoms** | 18 [10, 33] | 7 [4, 11] | < 0.001 |
| **Duration of isolation** | 16 [11, 25] | 7 [5, 12] | < 0.001 |
| **Productivity loss due to**<br>**isolation (JPY)** | 1424.3 [825.6, 2545.5] | 606.1 [297.0, 1090.9] | < 0.001 |
| **Quality of Life** | 0.89 [0.72, 0.97] | 0.96 [0.86, 0.99] | 0.001 |

ILI; influenza-like illness, USD; United States Dollar, JPY; Japanese Yen (1 USD = 110
JPY)
\*Results of Mann-Whitney U test
Numbers with square brackets represent median value and interquartile range

Figure 1 shows the probability density curve of duration of symptoms and isolation, productivity loss, and Quality of Life at the day of questionnaire survey.

**Figure 1.**
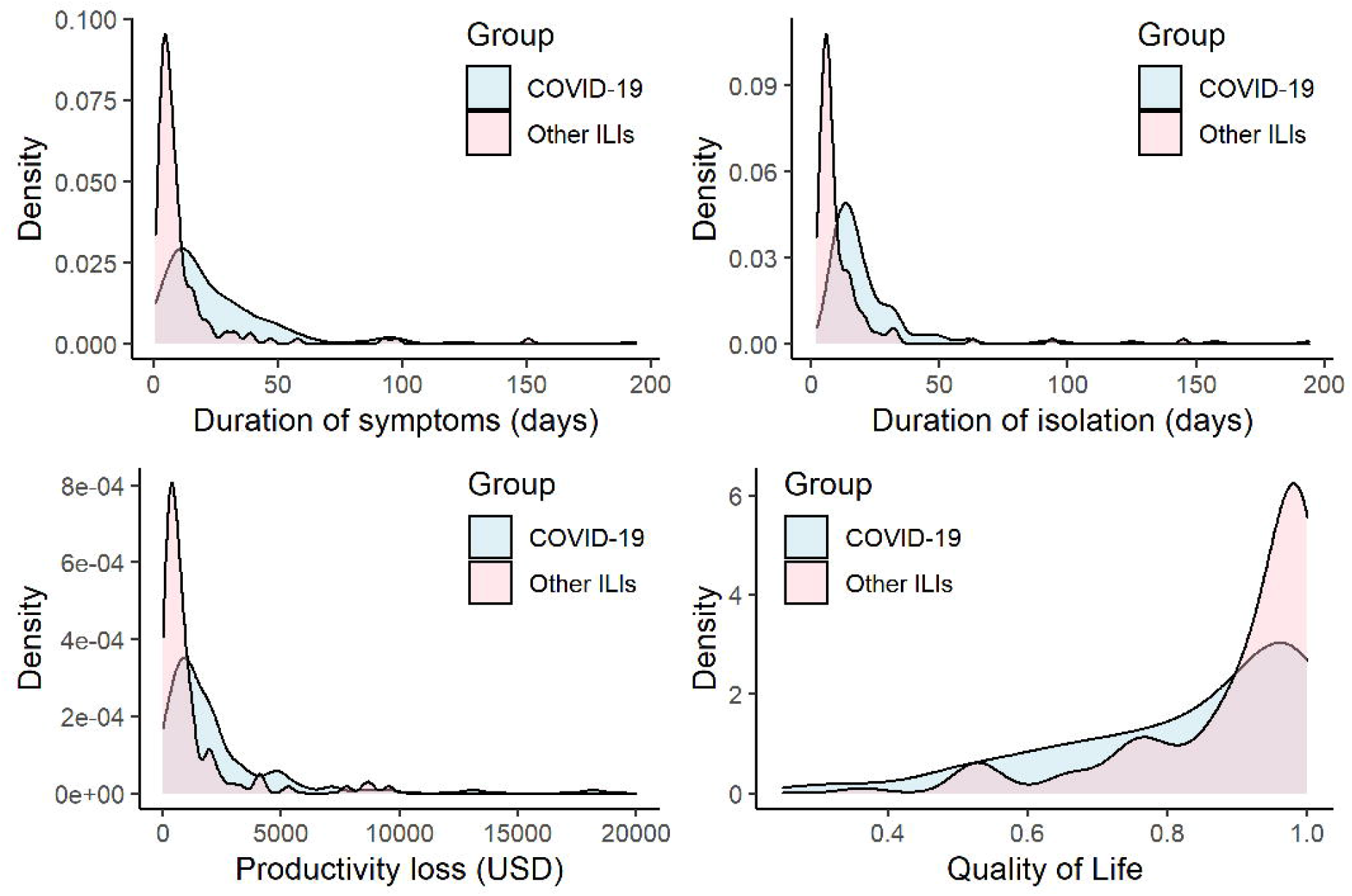
Probability density curve of duration of symptoms and isolation, productivity loss and Quality of life. Top left panel shows duration of symptoms. Top right panel shows duration of isolation. Bottom left panel shows productivity loss. Bottom right panel shows Quality of life. ILI; influenza-like illness, QOL; Quality of life Red area represents COVID-19 group and Blue area represents other ILIs group.

## Discussion

We showed the difference between COVID-19 and other ILIs in relation to the individual QOL and economic impact of isolation. As expected, COVID-19 imposes us heavier burden on our society than other ILIs such as seasonal influenza do in various aspects.

First, the longer duration of isolation brings us greater absenteeism. It is noteworthy that the median duration of isolation (16 days by the data after propensity score matching) was longer than 14 days. As described above, its duration in Japan was determined as 14 days at the early stage of the pandemic and changed to 10 days later by Ministry of Health, Labour and Welfare (MHLW) [15]. Japanese government also adopted the condition defined by WHO [16] that “plus at least 3 additional days without symptoms (including without fever and without respiratory symptoms)” then longer duration of symptoms bring us longer duration of isolation.

As for other ILIs group, the most frequent diagnosis was seasonal influenza (74.4%). It was probably due to the isolation policy specific to Japan [23]. Not MHLW but Ministry of Education, Culture, Sports, Science and Technology defined the isolation period due to seasonal influenza for school children based on School Health and Safety Act [24]. This law defines five-days isolation period for seasonal influenza with the day of symptom onset as day 0. It is a main cause of comparatively long isolation period for seasonal influenza in Japan [23]. Therefore, the burden caused by isolation due to seasonal influenza in Japan seems heavier than that in other countries, nevertheless, that caused by COVID-19 imposes a heavier burden.

As a result of longer duration of isolation, its productivity loss became larger in COVID-19 group than in other ILIs group. In addition, our results showed the productivity loss from patients’ perspective. If we consider societal perspective, productivity losses caused by other aspects might be another burden to our society. For instance, different from other ILIs, COVID-19 patients were often isolated (in case the patients have no risk factors of severe diseases and their cohabitants have any risk factors)[15] at not their own home but some hotels. Although their accommodation fee and healthcare professional personnel costs are covered by the government, the quantitative evaluation of these costs are not available.

QOL value is another point to be discussed. Even after we adjust participants’ background of two groups, there were significant differences in QOL value at the day of questionnaire survey between COVID-19 group and other ILIs group (0.89 and 0.96, respectively). This result might suggest that, even if patients were not aware of its symptoms, COVID-19 had long-term effect on their health status.

So-called “long COVID” or “post COVID syndrome” is not fully defined [25,26], however, one of the phenomena specific to COVID-19 compared with other ILIs and it will bring us further additional burden on our society. In our survey, more than a half (70/138, 50.7%) of participants who had COVID-19 infection answered that their symptom(s) lasts more than four weeks. Conversely, only 13 out of 164 (7.9%) in other ILIs group showed the duration of symptoms longer than four weeks. The proportion of patients who reported “long COVID” varies between studies [27,28], and our result (50.7%) is not substantially different from these findings. Considering this, even after its acute phase, COVID-19 may continue to have a negative impact on QOL and productivity, more frequently and longer than for other ILIs.

Our study includes several limitations. First, as our data were based on an online survey, it requires participants to have basic internet literacy. That is, participants of our survey might have more interest in their health status and better knowledge of basic computational skill than the general Japanese population do.

Second, as already discussed in this section, productivity loss was evaluated only from participants’ viewpoint, and no data are available about its societal perspective (accommodation fee, healthcare professional personnel, etc.). Third, QOL was assessed at the day of questionnaire survey and not assessed at the time they had symptoms due to COVID-19 or other ILIs because 15-D questionnaire does not set any recall period. More precise quantitative evaluation of productivity loss and health-related QOL will be a future challenge.

In conclusion, our results showed that COVID-19 imposes heavier burden on us than other ILIs do, not only its symptoms but also productivity loss due to its longer duration of isolation. These findings will be helpful for healthcare policy makers to determine the appropriate duration of isolation. “long COVID” might be a common phenomenon which bring us an additional burden, then quantitative evaluation of the burden of “long COVID” would be one of future challenges as well as more precise assessment of productivity loss and QOL value.

## Data Availability

The data that support the findings of this study are available upon request to the corresponding author. The data are not publicly available due to privacy or ethical restrictions.

## Acknowledgment

We thank all the people who participated in our survey.

## Author contributions

Shinya Tsuzuki: Conceptualization, Funding Acquisition, Data Curation, Formal Analysis, Methodology, Visualization, Project Administration, Original Draft Preparation. Norio Ohmagari: Funding Acquisition, Project Administration, Supervision, Review and Editing. Philippe Beutels: Conceptualization, Methodology, Supervision, Project Administration, Review & Editing.

## Ethics approval

This study was reviewed and approved by the Ethics Committee of the Center Hospital of the NCGM (NCGM-G-004001-01).

## Patient consent

Written informed consent was obtained by each participant before starting the survey through an electronic form.

## Permission to reproduce material from other sources

Not applicable.

